# Women’s Preferences for and Experiences with Contraceptive Side Effect Counseling and Management: A Qualitative Study from Ethiopia and Kenya

**DOI:** 10.1101/2025.11.24.25340915

**Authors:** Lauren Suchman, Beth Phillips, Serah Gitome, Hailemichael Bizuneh, Hanna Feleke, Zachary Kwena, Sarah Okumu, Pauline Wekesa, Elizabeth Bukusi, Kristen Kirksey, Kelsey Holt, Jenny Liu, Ewenat Gebrehanna

## Abstract

**Objective:** While not inherently negative, the magnitude of contraceptive non-use attributed to side effects globally merits concerted attention to ensure women receive quality side effect counseling and support. Evidence shows comprehensive contraceptive counseling can help women navigate side effects, yet survey data reveal consistent gaps in side effect counseling quality. To elucidate women’s lived experiences with side effect counseling and management, we examined how women in Kenya and Ethiopia manage contraceptive side effects with and without help from healthcare providers, and what their preferences are for support.

**Methods:** We conducted in-depth interviews with sexually active women of childbearing age (N=83) in Nairobi and Kisumu, Kenya, and Addis Ababa and Amhara region, Ethiopia. Participants were asked about their contraceptive experiences, including side effects, how they managed side effects, what support they received, and their preferred approaches for receiving support when experiencing side effects. We utilized a modified grounded theory approach for data collection and analysis.

**Results:** We found gaps in reported counseling quality among women in our sample, and a desire for more comprehensive counseling on side effects during initial and follow-up visits. After experiencing side effects, most participants described enduring and self-managing with over-the-counter remedies, and seeking clinical advice only when they worried about how their contraceptive method was affecting their overall health. Follow-up services often addressed participants’ immediate needs, but rarely offered additional information or comprehensive counseling. A few participants were denied a desired contraceptive device removal or shamed for not using contraception.

**Conclusion:** We identified a need to improve anticipatory contraceptive side effects counseling and proactive follow-up approaches that support women to choose methods they prefer and then navigate their use or switching. Such counseling would help health systems become more responsive to patient needs in contexts where women’s suffering is often considered acceptable, rather than cause for intervention.

## INTRODUCTION

Many women are deterred from using contraception due to fear of side effects [1–4] or discontinue contraception when side effects are uncomfortable or difficult to manage [5–8]. In Ethiopia, 16% of women who are not trying to get pregnant cite health concerns, including side effects, as a reason for not using contraception [9], while a recent study in Kenya found that 61% of participants who reported a reason for switching contraceptive methods cited side effects as their main concern [10]. Notably, contraceptive use in and of itself is not inherently good. Some women may make an informed decision that the benefit of preventing pregnancy does not outweigh the potential side effects or social consequences associated with using contraception [12]. However, nonuse or discontinuation of contraception due to lack of high-quality information or support for managing side effects represent missed opportunities to better enable women to manage fertility in line with their preferences. Indeed, contraceptive counseling that includes information on potential side effects has been associated with continued use of family planning among women wanting to prevent pregnancy [13]. Further, evidence shows that when providers counsel specifically on side effects, clients are significantly more likely to use contraception and to seek care if they experience side effects [14–16].

World Health Organization family planning guidelines and other contraception quality of care frameworks emphasize the importance of discussing side effects during method selection and providing follow-up support to help women manage side effects or switch methods if so desired [17,18]. Yet, in sub-Saharan Africa (SSA), poorly funded health systems, inadequate infrastructure, and/or obligations to meet numeric targets often have deleterious effects on the quality of reproductive healthcare in general, and contraceptive counseling specifically [19–21]. These structural issues create a context in which providers often have limited capacity, motivation, or opportunity to deliver comprehensive and effective counseling, [22,23] including counseling on side effect management [24]. the proportion of women who receive information about side effects during contraceptive counseling, as measured by the Method Information Index (MII) [25], is consistently low globally [26–29]; 59% and over 80% of women do not receive comprehensive counseling in Kenya and Ethiopia respectively [30,31] and evidence from Ethiopia shows a significant decline in MII index scores from 2015 – 2019 [28].

A small body of qualitative research from sub-Saharan Africa on experiences with side effect counseling expands upon MII Index estimates. For example, Yirgu et al. found that women in Ethiopia felt providers were giving them inaccurate information about side effects associated with short-term contraceptive methods to encourage them to use long-acting methods instead [32]. Senderowicz found that providers in an undisclosed sub-Saharan African country withheld information about side effects during counseling sessions for the same reason [33]. However, beyond experiences with side effects counseling during method selection, little is known about women’s lived experiences navigating side effects when they occur and their preferences for follow-up support. Alarmingly, some research has shown that rather than being supported to manage side effects or switch methods when they become intolerable, women are, in some cases, denied removals (of implants or intra-uterine devices) [34–37].

To add to the literature on contraceptive side effect experiences, we sought to elucidate how women manage contraceptive side effects with and without the help of healthcare providers, and what their preferences are for side effect counseling and management. We conducted 83 semi-structured in-depth interviews with sexually active women of reproductive age in Ethiopia and Kenya. Our findings have implications for policymakers, program developers, and implementers seeking to improve contraceptive counseling and follow-up services in these countries, with particular attention to side effects counseling and management.

## METHODS

We conducted a cross-sectional qualitative study as part of the Innovations in Counseling and Follow-up (ICAF) project. ICAF’s main objectives are to: 1) understand women’s preferences around contraceptive side effects; 2) develop and test interventions to improve contraceptive counseling and follow-up care for the navigation of bleeding-related and other side effects. We chose Ethiopia and Kenya for the ICAF project to represent different sociocultural and health system contexts in East Africa.

### Study sample and recruitment

Data were collected in Addis Ababa and the North Shoa Zone of the Amhara region in Ethiopia, and Nairobi and Kisumu counties in Kenya. Addis Ababa has the highest contraceptive prevalence in Ethiopia [38] and contraceptive services are technically free to married women in public health facilities, although some have reported unauthorized fees [39]. In contrast, North Shoa is an agrarian community with limited health service access and options for contraceptive care. Nairobi and Kisumu are both cosmopolitan cities with large populations of women who are low socioeconomic status and face multiple barriers to accessing contraceptives [14,40]. Nairobi is a large urban center that draws residents from across Kenya, while Kisumu is unique due to its proximity to rural populations. Both cities have a mix of public and private contraceptive service providers and these services are technically free in public health facilities, although this is not always the case in practice [41].

From April through May 2023, we recruited a sample of N=83 (n=43 in Ethiopia, n=40 in Kenya) sexually active women between the ages of 15-45 years. In both study countries, we took a purposive sampling approach, sampling for contraceptive use (those who had ever used contraception and those who had not), age (15-19 years, 20-45 years), and marital status. Participants were recruited primarily in public health facilities with some recruitment from communities. Women recruited in clinics were approached by the study’s data collectors, assessed for eligibility and enrolled in the study as they visited family planning services. In both study countries, we gave local community health workers our study eligibility criteria and asked them to refer potential participants for screening and possible enrolment. After receiving women’s permission, community health workers shared their contact information with the study team. Data collectors then contacted potential participants via phone to assess eligibility and enroll women who met the eligibility criteria and gave their informed consent to participate.

### Data collection

Research Assistants (RAs) were all women and fluent in the relevant local languages (Amharic in Ethiopia, Swahili and Dholuo in Kenya) and English. We conducted semi-structured in-depth interviews to gain a deeper understanding of women’s experiences with and preferences regarding contraceptive side effects, their management, and side effects counseling. Senior researchers with qualitative experience, which were study leads from each participating institution, drafted the interview guides, then reviewed before the guides were field tested and data collection began. As data collection progressed, the study team added and refined some of the probes within what was allowable by the appropriate ethical review boards. We also fielded a short demographic survey with each interview. Interviews took place in a location of the participant’s choosing (e.g., the grounds of the health facility or outdoor spaces removed from foot traffic), and RAs prioritized auditory privacy when finalizing the location. In Ethiopia, we obtained verbal informed consent from participants before conducting interviews. RAs read the consent document aloud to potential participants, requested that potential participants give verbal consent, and then the RAs signed the form themselves to document that consent was obtained. In Kenya, the Ras obtained written assent for girls aged 15-17 and written consent for women aged 18+. We did not obtain parental consent for girls aged 15-17 to participate, as girls might not be willing to disclose their contraceptive use to parents due to fears of potential consequences. After participants gave informed consent, interviews were audio recorded and lasted 1-2 hours. Audio files were uploaded to an encrypted server before being translated and transcribed for analysis. In addition, RAs used a note-taking template to record de-identified field notes, which they shared with the research team to guide probing in subsequent interviews and similarly saved to an encrypted server.

### Data analysis

Following a process previously established by our team, we built on Charmaz’s modified grounded theory approach [42] by conducting ongoing team-based rapid analysis throughout the data collection period [43]. This involved RAs completing memo templates after each interview that summarized the data relevant to our main research questions. Members of each study country team reviewed newly available transcripts weekly as they were completed over time and met to discuss emergent themes. Weekly meetings were complemented by biweekly cross-country team meetings to discuss similarities and differences across study countries and agree on areas to probe further in subsequent interviews. This approach also allowed us to adjust sampling targets for remaining interviews to ensure we reached thematic saturation, and to iteratively adjust interview guides (within parameters allowed by ethics review boards). Regular meetings between the research teams ensured ongoing alignment between the tools while allowing for small adjustments to bolster local relevance.

Interview audio recordings were transcribed by the RAs in Ethiopia and by a team of independent transcriptionists in Kenya. In both study countries, members of the research team uploaded the final transcripts to Dedoose qualitative analysis software for analysis. Using a codebook that several team members developed—both deductively drawing on key research questions and inductively drawing on findings from the rapid analysis—a team of 12 experienced coders from Ethiopia, Kenya, and the United States then participated in an inter-coder reliability process. This process consisted of coding the same transcript, getting an intercoder reliability score through Dedoose, and then meeting as a group to discuss discrepancies in coding. Coders from each study country team then divided and individually coded a set of transcripts from their country. Members of the UCSF team were assigned to code transcripts from each country as needed. All coders participated in weekly analysis meetings to continue discussing emerging themes and refine the codebook. Researchers from all teams then generated reports for codes relevant to side effect experience and management (e.g., “self-management side effects” and “experience – contraceptive counseling”), drafted code summaries, and developed themes through memo-ing and cross-team discussion.

### Ethical approvals

In Kenya, the study team obtained ethical approvals from the Maseno University Scientific Ethics Review Committee (MUSERC) (MUSERC/1171/22) and the National Commission of Science and Technology Innovation (NACOSTI) (NACOSTI/P/24/33360). In Ethiopia, we obtained approval from the Institutional Review Board (IRB) of Saint Paul’s Hospital Millennium Medical College (SPHMMC) (PM23/487). In addition, we received ethical approval from the UCSF IRB (22-37961).

## RESULTS

Our sample consisted of 83 women. As shown in Table 1, most participants were contraceptive users (67%) aged 20-45 years (59%) with at least one child (78%). While most of our sample (88%) in Ethiopia was married, the sample in Kenya was more even between married (45%) and unmarried (48%) women.

**Table 1.**
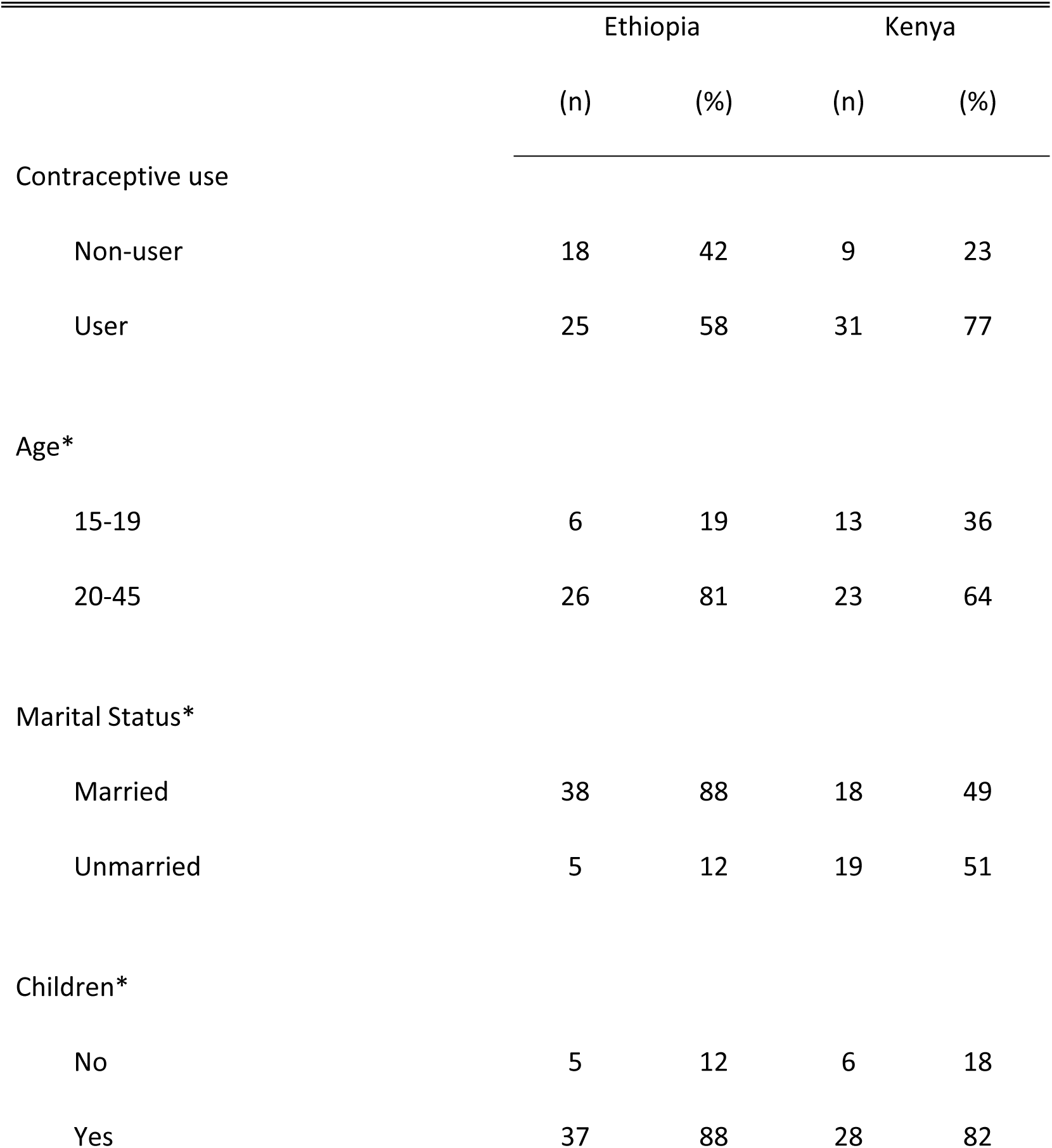
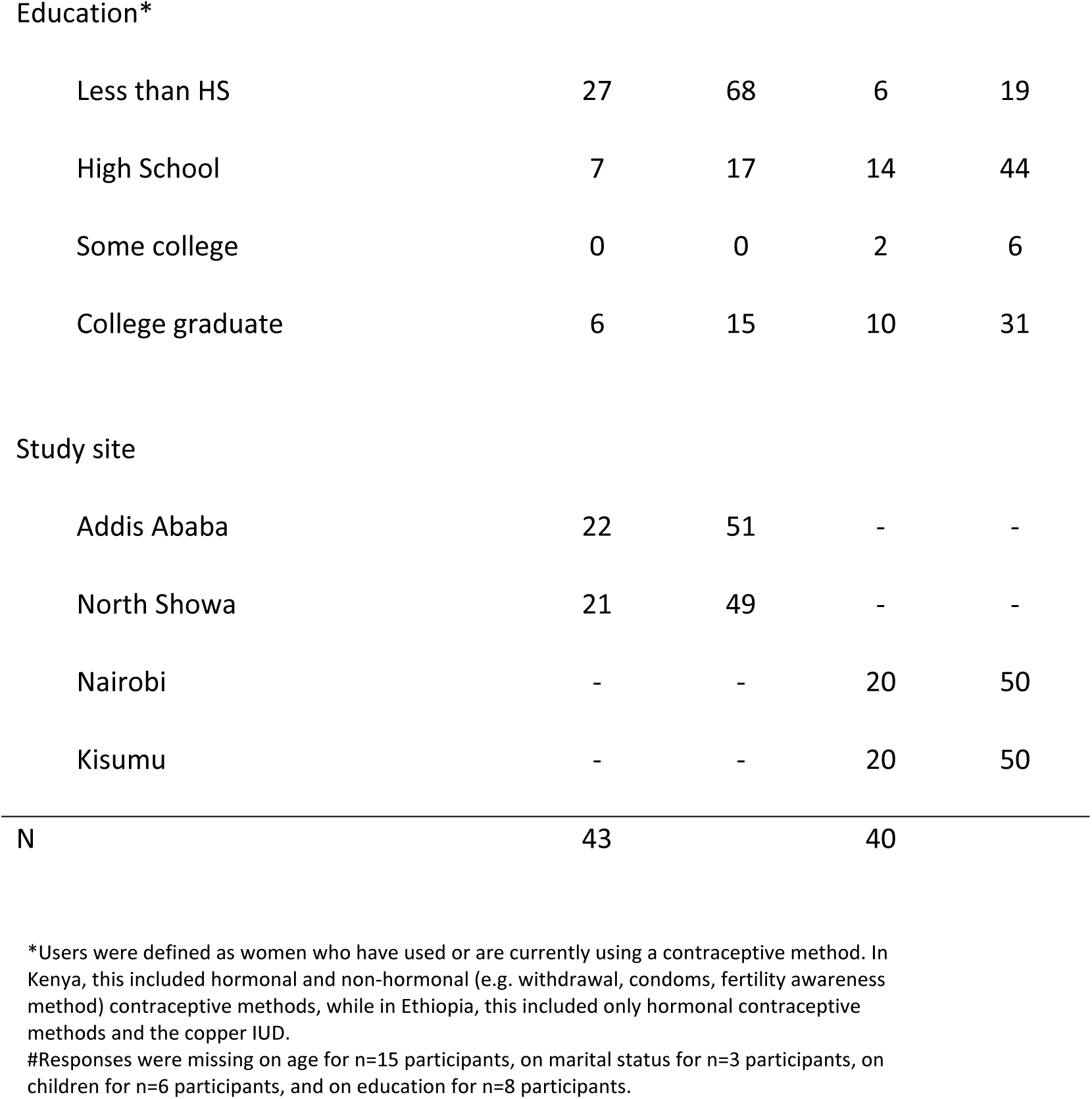
Descriptive characteristics of sample (N=83)

Further, 60% of the women interviewed in Ethiopia had less than a high school education, while most of the participants in Kenya had completed high school or above.

### Participants assessed implications for their health and well-being after first experiencing side effects, then used a range of strategies to cope

Women in our sample described experiencing a range of side effects with their contraceptive use. These ranged from heavy bleeding and frequent periods to amenorrhea, weight gain to weight loss, and included nausea, headaches, cramps, body aches, and loss of appetite. Many who had never used contraception described a fear of experiencing and managing side effects as a primary reason why they had not pursued contraceptive use.

Reflecting on why they chose a particular contraceptive method, most women in our sample said they expected some side effects when starting a new method, but did not think proactively about how the side effects associated with different methods might affect their daily life while making their initial choice. Instead, many said that they started a new method and then had to take time to determine whether a method was “suitable.” For most participants, “suitability” indicated that they were able to maintain their health (e.g., they did not suffer from anemia) while using a particular method.

> *Ok. This is the thing. You see, for the contraceptive, when you start using it, you use it, and **after using it, it’s when you will know if your body suits it or not**.* (contraceptive user, 20-45 years, unmarried, Kenya)

Except in cases where they believed side effects had negatively affected their health or ability to conduct daily activities, many participants accepted contraceptive side effects as bothersome aspects of their everyday lives, which they should manage themselves because it was their fate as women to suffer.

> *I haven’t consulted anyone’s saying, “I’m sick, it’s burning.” I don’t know, it’s my problem.*

> (contraceptive user, age unknown, married with children, Ethiopia).

> *I just feared that I had only a few complications and I also got ideas from my friends saying that it is normal. Yeah, that it’s normal.* (contraceptive user, 15-19 years, married with children, Kenya)

Indeed, many women felt they had no choice but to continue using contraception regardless of side effects experienced, because preventing pregnancy was a higher priority than their own discomfort.

> *I started having skin color changes. I didn’t have hyperpigmented skin before. I also have loss of appetite; when I eat, I would feel full after two bites. I am using the contraceptive because **it is better to feel discomfort than have a child and suffer**.* (contraceptive user, age unknown, married with children, Ethiopia)

To manage side effects, women in both study countries described taking herbal supplements or vitamins, or eating iron-rich foods (i.e., liver) when they had heavy bleeding and worried about anemia. Many also took over-the-counter painkillers, such as paracetamol, to ease cramps or headaches. Some women in Ethiopia mentioned hearing from others that they should drink warm water or tea to help with pain, and similarly, some had heard that extra fluids could increase menstrual flow when it was unusually light due to contraceptive use.

> *When we are on [menstrual] period, we are usually told to use hot beverages because it will reduce the pain.* (contraceptive user, 20-45 years, married with children, Ethiopia)

Study participants in Kenya also mentioned carrying extra sanitary pads with them or wearing dark clothing to mask overflow in cases where their contraceptive caused unusually heavy menses.

> *Right now, I usually carry emergency pads. So, I don’t walk without pads.* (contraceptive user, age unknown, married with children, Kenya)

When experiencing side effects, many women in our study reported consulting with female family members or peers to determine if their side effects were normal or required a visit to a healthcare provider. In these cases, knowing that a person they trusted had gone through something similar helped reassure women that certain side effects, such as weight gain or irregular menstruation, were not a cause for concern.

> *Participant: **Me and my sisters talk about it.** Since they’re using contraception as well, we check on each other. They’d often tell me that they used to be like me, but now they’re doing well. Interviewer: What was their response when you told them about your changes?*

> *Participant: I told them that my period comes every 15 days, and they said that theirs was the same as well. They told me that it only happens until I get used to the contraception. **So, uh, I dropped that concern afterwards**.* (contraceptive user, 20-45 years, married with children, Ethiopia)

However, if contraceptive side effects were too uncomfortable, difficult to manage, or caused women to worry about their health, participants in both countries commonly reported that they switched methods or discontinued contraceptive use altogether.

> *I switch to this [method] after knowing that the implant is not compatible with me.*

> (contraceptive user, 20-45 years, married with children, Ethiopia)

> ***It caused me problems health-wise****. I had low blood levels, I couldn’t stand, eat, my health deteriorated, and so I had to stop.* (contraceptive user, age unknown, married with children, Kenya)

### Clients returning for follow-up care at a health facility tended to rely on providers to assess method suitability; providers often addressed clients’ concerns but did not offer additional counseling on other method options and their side effects

Several participants in both study countries described going to a healthcare provider when they experienced side effects that worried them and tended to rely on their provider to determine whether their current contraceptive method was suitable or not. This was especially common among women who experienced either too much (more than once per month or increased volume) or too little (missing their menses for more than two months) bleeding, or who thought their hormones were imbalanced because of their contraceptive use.

> *I mean normally your period might come and stay for days then it will stop, but this one it flows continuously. It kept flowing for almost two months. Even it makes me to lose my balance. **When I went [to the health center] they said, “this might happen on some women, it is not suitable for you”** and they changed it to injectable.* (contraceptive user, 20-45 years, married with children, Ethiopia)

> *It’s the doctor who told me to use this and this, he’s still the same person who will give me advice how to get out of this. So, me I believe **when it comes to health, the right person to deal with is a doctor**.* (contraceptive user, 20-45 years, unmarried with children, Kenya)

Women in our sample reported that, during most of these visits, providers listened to participants’ concerns about the side effects they were experiencing and either recommended a solution, such as pills to control bleeding, or suggested that the participant should wait longer for her body to adjust to the method she had chosen. However, in Ethiopia, at least one participant confirmed an experience with a provider who refused to remove a contraceptive method that was causing uncomfortable side effects because the provider was concerned that other women would then come in and also ask for removal.

> *Then we went to the health center and asked for removal. Then the health providers said that they wouldn’t remove it in such a short time. And **said that I’d rile up and bring other women if they let me**. They said that it wouldn’t be removed before its due date.* (contraceptive user, age unknown, married with children, Ethiopia)

Many participants in our study said that they took the provider’s advice with few questions. Those who felt like their provider had not fully addressed their concerns and had other options for contraceptive care in their area sought a second opinion.

> *That’s when I went to the second doctor who gave me the pills [to manage heavy bleeding related to injectable contraceptive use].* (contraceptive user, 20-45 years, married with children, Kenya)

Participants reported they were satisfied with follow-up care for side effects when they received treatments that reduced side effect severity, they felt the provider listened to their concerns and provided actionable advice, or the provider helped them get rid of their side effects by switching to a new contraceptive method. However, these visits often did not involve discussion about why women were experiencing side effects or opportunities to learn about the range of other methods available.

> *Participant: When I had faced total cessation of menses I went [to the health facility] and consulted them and they gave me support.*

> *Interviewer: Ok, how much were you satisfied from the care that you said I get from them?*

> *Participant: I felt very happy. I mean, when I had faced total cessation of menses they prescribed me to take a pill. Then after I took the pill it was corrected and flows properly. When I saw this I was very happy.* (contraceptive user, 20-45 years, married with no children, Ethiopia)

> *When I went [to the health facility] and they noticed that I have changed, they asked me about it and told me that’s the way those things react with people […] They didn’t tell me anything much, they just advised me to continue using the method and said I would stand a risk of getting pregnant if I stopped using it.* (contraceptive user, age unknown, married with children, Kenya)

Notably, providers at some health facilities followed up with clients proactively over the phone to see how they were doing with their new contraceptive method. Participants who were interviewed at a private health facility in Kisumu, Kenya, reported that this proactive follow-up gave them an opportunity to discuss side effects they were experiencing and ask the provider questions.

> *Participant: The support they have given me is to just have a follow-up. **They have been following up on me**, at least they have been making some calls in the course of the months before I go for my refills, before I go for another clinic day for contraceptive.*

> *Interviewer: So, what do they call you to tell you?*

> *Participant: To find out if I am okay, if I have any problem or any challenge, or if Depo is taking me negatively*

> *Interviewer: Did this support or care sufficiently address your needs?*

> *Participant: Yes, it has been addressing, at least it has made it much easier, I am not walking to go to the clinic to ask questions. At least it is just one call away, we can just communicate through the phone. (contraceptive user, 20-45 years, married with children, Kenya)*

> *After missing my menses, when I was going to get the pills, at least the hospital was calling me through their customer care number and they were following up to see how I was doing.* (contraceptive user, 15-19 years, married with children, Kenya)

Conversely, one participant in Ethiopia noted that her healthcare provider proactively shamed her when she did not follow the provider’s advice.

> *Interviewer: Earlier you said that you had come here to get the three-month method. Why did you use the three-month one?*

> *Participant: Well, what can I do when she [health care provider] told me to use it? Interviewer: What did she say to you?*

> *Participant: She said I should not have another child. And she told me to come in and get the three-month method. But when I got checked, they told me that my blood had dried out [low blood pressure] and that the injections were not suitable for me. **So, I left but then she called me over the phone and yelled at me for not getting the method**.* (never user of hormonal contraception or copper IUD, 20-45 years, married with children, Ethiopia)

### Participants wished initial counseling sessions had included more information about a range of contraceptive options and their potential side effects, along with opportunities to ask questions and proactively learn about side effect management

When asked about experiences with contraceptive counseling, most study participants lamented that their provider offered limited information on side effects when they were first counseled on method choice and did not offer opportunities for the woman to ask questions. Experiencing side effects then felt surprising or confusing for some.

> *I just told [the provider] that I need a contraceptive but I only knew about Depo and pills at that time. So, I told the doctor that I don’t want pills, but I want the other one but not pills. **But he didn’t explain for me that there is Depo, there is coil, there is what what**. So, he told me that I can inject you this one for three months. Then I just say okay how much is it, and then he told me the amount. And I just decided to put it. (contraceptive user, 15-19 years, married with children, Kenya)*

> *Now, when I got sick because of the contraceptive I said to myself, why they didn’t tell me that it will bring this.* (contraceptive user, 20-45 years, unmarried with no children, Ethiopia)

One notable exception was a participant in Kenya who reported an especially positive experience with a community health promoter (CHP) she encountered through an outreach event sponsored by a local NGO. This participant felt the CHP was motivated to provide comprehensive counseling to help her organization achieve its program goals:

> *Participant: [My experience] was good because the CHPs were explaining those things very well for us. Side effects and everything because they wanted people to go and get the services so that their outreach day could be successful and that is why they were giving a lot of information about it.*

> *Interviewer: Okay, so what other information that you were given, one you have said it was about the side effects, anything else?*

> *Participant: Yes, the side effects, then the good side of it; you could remove it and you will not take long before conceiving if you want to.* (contraceptive user, 20-45 years, married with children, Kenya)

In some cases, Kenyan participants felt that providers intentionally withheld information on side effects to encourage contraceptive uptake, while others believed their provider was too busy to engage with them in an extensive discussion about contraception at the time of their counseling visit.

> *Interviewer: Did she talk to you maybe about feeling tired?*

> *Participant: **No, they do not talk about those because it would prevent you from going for the services**. So, things like headache, obesity, they do not talk about (laughter). They will just talk about normal things. They can’t tell you that if you go for this method then you will be so tired, no.* (contraceptive user, 20-45 years, married with children, Kenya)

> ***There was no time, she just rushed the information ‘cause she was busy*** *and told me that [the contraceptive method] would be put [in] when I came back for baby’s clinic.* (never used contraception, 15-19 years, married with children, Kenya)

At the same time, some participants reported that they had to accomplish several tasks while visiting the clinic, such as getting their child vaccinated, or needed to return home for childcare. This made women themselves rush through their appointment without asking questions.

> *My child was crying alone so I went out to go to him. I took him and I went. That’s it.*

> *(contraceptive user, 20-45 years, married with children, Ethiopia)*

> *I also wanted to go and breastfeed the baby ‘cause he was in nursery. So, I did not ask questions.* (never used contraception,15-19 years, married with children, Kenya)

Typically, though, participants noted that their providers had recommended methods or offered a limited range of choices based on duration of effectiveness (i.e., how long a woman would need the method to work before she could return to the clinic or hoped to get pregnant) with less attention paid to how a woman would otherwise experience the method on a day-to-day basis (i.e., side effects).

> *I came here and consulted them. I told them that I needed family planning. **They asked me how many years I was giving them**. They told me about the five years and the other methods. They told me about everything. So, I decided on five years because I didn’t want to nag myself [to seek contraceptive refills more often].* (contraceptive user, 20-45 years, married with children, Ethiopia)

> *When I talked to the nurse, the nurse said, when you want to get another kid, you can calculate the time when you want to get another kid then you will be okay with it, but the nurse was telling me; I choose between coil and [implant]. So, I saw it like staying with coil for 10 years. It is just hectic for me and that is why I settled with [the implant].* (contraceptive user, 15-19 years, married with children, Kenya)

Even if they were confused by the information provided in this first session or had additional questions, some felt nervous asking questions in a healthcare setting, did not know what kinds of questions to ask in a follow-up conversation, or felt they did not have the right to ask. In such situations, some participants suggested it was up to the provider to prompt conversation and create a safe space for open discussion.

> ***There is nothing, what would I ask****? If there was something I wanted to ask, I would have asked, but there was nothing I wanted to ask, and they didn’t ask me anything, I didn’t ask them anything.* (contraceptive user, age unknown, married with children, Ethiopia)

> *Interviewer: Okay, so what do you think should be done so that you feel you are comfortable with someone to [discuss] these bed issues [issues related to sexual intimacy]?*

> *Participant: What should be done is just the provider to talk it out before the client raises it up. Yeah, so it would give you an easy time; that space of talking about many things. **Now if the providers are not talking about it, they are not willing to raise it up, then it becomes hard for you as a client to ask.** However much you feel like asking such questions, but there is a pulling force that holds you from asking about it.* (contraceptive user, 20-45 years, married with children, Kenya)

In addition to offering opportunities to ask questions, a few women noted that they wanted to not only learn about potential side effects but also to discuss specific management strategies with their providers.

> *I wanted them to tell me about the reasons why the side effects occur, what to do when I feel pain, and how to prevent losing weight.* (contraceptive user, age unknown, married with children, Ethiopia)

## DISCUSSION

Our interview findings from women in two East African countries help elucidate how women manage contraceptive side effects with and without the help of healthcare providers, and what their preferences are for side effect counseling and management. We found gaps in counseling quality experienced by women in our sample, and a desire for more comprehensive counseling on side effects associated with different method options during method selection and in follow-up visits for bothersome side effects.

Women in our study often endured side effects—using a variety of self-care management techniques—and sought clinical advice only when side effects threatened their health or quality of life. During follow-up care, some received welcome support for management of side effects while a minority was denied a desired contraceptive device removal or shamed for not using contraception.

Overall, our findings lend additional evidence for poor quality of counseling related to contraceptive side effects and, despite longstanding quality of care guidelines in the family planning field [44] that emphasize the need for continuity of care, our findings corroborate other recent calls for a need for more attention to follow-up support for side effect management [34]. As some participants in our research suspected, other studies from SSA have documented providers limiting or manipulating information on side effects in order to encourage, or even coerce, women into accepting certain methods [32,33]. Similarly, evidence from the U.S. shows that providers neglect to discuss or de-emphasize negative side effects when helping women to make a decision about contraception [45,46]. Building on this suspicion that they were not receiving comprehensive counseling, it was clear that participants in our study wanted more information on side effects during contraceptive counseling sessions, similar to findings from myriad geographies [47,48].

Some tools to improve contraceptive counseling have shown promise when designed to enhance counseling specifically related to side effects [49], while others have helped women feel more comfortable asking questions in counseling sessions [50]. Yet, reviews of contraceptive counseling tools—including provider job aids and digital tools for women—reveal major gaps in side effect decision support and anticipatory side effect management guidance [51]. While this gap in existing tools highlights an opportunity for innovation, it is important to note that tools such as counseling guides and decision aids often show mixed effectiveness in real-world implementation settings, where providers may not have the resources or desire to regularly use decision aids during clinical encounters [52,53].

Indeed, our findings suggest that providing additional information is not sufficient to help women make choices that are aligned with their values and priorities. Counseling providers should also give clients the opportunity to ask questions and engage women in discussion about how contraceptive side effects might materially affect their health and daily lives. Thus, complementary approaches that do not rely solely on improving providers’ counseling are needed to enhance informed choice. As noted by one of our participants, community health volunteers (CHVs) can be effective for increasing the reach of the formal health system [54,55], particularly among populations that are hardly reached with contraceptive services [56]. However, the literature assessing the quality of CHV services has shown wide variability and suggests that performance is highly context-specific [57]. Further, to our knowledge, these programs have not been evaluated for quality of contraceptive counseling.

Additionally, it is important to highlight that many women in our sample faced challenges advocating for themselves and asking questions during both initial and follow-up contraceptive counseling sessions. While some participants felt empowered to engage in helpful discussions with their providers, a number of participants reported that they weren’t always sure they could trust their provider to give comprehensive information, and many felt nervous to ask follow-up questions. A minority described mistreatment from providers, either denying removal of contraceptive devices or shaming them for not using recommended contraception. These findings are in line with an extensive body of literature on mistreatment, including physical and verbal abuse, in reproductive healthcare settings in Kenya and

Ethiopia, as well as across SSA [58–60]. However, much of this mistreatment may be the result of systemic factors beyond providers’ control. These include being overburdened by workload [61], the generally stressful nature of their work [62], and, as our team has previously documented in Ethiopia, pressure to meet performance targets [21]. However, we note an extensive body of literature demonstrating that, in many patriarchal healthcare systems, women’s suffering is considered acceptable to an extent that is not acceptable for other populations [63,64]. Thus, in order to provide quality care [18], we contend that it is the duty of the health system to implement strategies that anticipate and proactively manage women’s suffering related to contraceptive side effects even when they themselves do not demand such support.

Accessible follow-up support for women experiencing side effects has the potential to help women decide whether to switch methods and what method to switch to or manage side effects if they decide to continue the method, yet virtually no evidence-based approaches for proactive follow-up counseling exist. Our finding that some participants in Kenya received a follow-up call from a private facility after initiating a new hormonal method is a promising example of simple and effective approaches to proactive follow-up care for side effects. Additionally, we found that many women use self-care techniques to manage side effects on their own; health system interventions to improve follow-up care could consider integrating acknowledgement of these self-care techniques with the broader set of medical management options, such as non-steroidal anti-inflammatory drugs or oral contraceptive pills for irregular or heavy bleeding. In response to findings from this study, our team has developed the *Mittin* (Ethiopia)/*FP Chap Chap* (Kenya) intervention. *Mittin/FP Chap Chap* is meant to improve contraceptive counseling and follow-up care for the navigation of bleeding-related and other side effects. The intervention consists of 1) enhanced clinic-based counseling via a counseling tool that improves upon the current standard of care for anticipatory side effect counseling and client education materials; 2) follow-up care through a call center where trained operators both provide proactive follow-up support and receive calls for on-demand side effects counseling. We conducted a three-month feasibility study of the *Mittin* intervention at one public health clinic in Addis Ababa and plan to test the intervention’s effectiveness in various contexts in future studies.

### Limitations

Our data come from a cross-sectional qualitative sample from two East African countries. Although these data were purposively collected to answer questions related to experiencing and managing contraceptive side effects, we collected data in cities that were urban or semi-urban in each country with some data collection in agrarian areas of Ethiopia due to research feasibility considerations. These constraints limit relevance of our findings for more rural areas of East Africa. Further, when we asked about participants’ experiences with contraceptive counseling, we may have introduced recall bias by asking about past experience because we did not set any time limit on the prior experiences individuals could comment on. Finally, we draw conclusions about the client-provider relationship based on reports only from clients themselves with no representation of providers in the dataset.

## CONCLUSION

Findings from our large qualitative study of women in two East African countries highlight gaps in anticipatory contraceptive side effects counseling as well as follow-up support once women experience side effects. Enhanced contraceptive counseling and follow-up approaches that center support for side effects, such as the *Mittin/FP Chap Chap* intervention developed by our team, could help facilitate empowered decision-making and women’s ability to manage side effects or switch methods in line with their preferences. Improving quality of contraceptive counseling and follow-up in this way can help people make informed decisions and navigate contraceptive use when they desire pregnancy prevention.

## Data Availability

All relevant data are within the manuscript. Supporting Information files (in-depth interview guides and codebook) may be found at 10.5281/zenodo.17633781.

10.5281/zenodo.17633781

## Acknowledgments

We wish to thank the research participants for their generosity of time and spirit. We would also like to thank Negin Zahedikia, Gladys Odhiambo, Titus Arunga, Vallery Obure, Mercelline Odhiambo, JaneRose Kweyu, and Sophie Ramirez for their contributions to the study.

## Funding

This work was supported by the Gates Foundation (INV-044992).

## Notes

### Competing Interest Statement

The authors have declared no competing interest.

